# UNDERSTANDING THE DETERMINANTS OF FLOURISHING AMONG ADOLESCENTS OF MAYUGE AND MPIGI DISTRICTS IN UGANDA: LINKING PHYSICAL, MENTAL, CULTURAL, ECONOMIC, AND SOCIAL DIMENSIONS THROUGH A GENDER-TRANSFORMATIVE LENS

**DOI:** 10.1101/2024.01.04.24300867

**Authors:** Benson Mutuku Muthama, Carol Henry, Judy White, Jessica Lieffers, Susan Fowler-Kerry, Daniel Kikulwe, Rebecca Tiessen, Lydia Kibiri, Benjamin Lough, David Musoke, Asrat Dibaba Tolossa

**Affiliations:** Department College of Pharmacy and Nutrition, University of Saskatchewan; University of Regina; Assistant Professor, York University; Faculty of Social Sciences, University of Ottawa; Makerere University, Kampala, Uganda; Director of Social Innovation at the Gies College of BusinessContact, University of Illinois; Department of Disease Control and Environmental Health, Makerere University; Medical Doctor and Public Health Specialist. World vision Canada

**Keywords:** Determinants, Flourishing, Adolescent, Gender, Gender-transformative, Gender Based Violence, Sexual Gender Based Violence

## Abstract

The study sought to engage adolescents in Uganda to learn about their perspectives on the factors that enable or hinder their flourishing. This cross-sectional study was conducted in Mpigi and Mayuge districts in Uganda, using a survey with adolescent boys and girls aged 14-19 years. The survey examined seven aspects of human flourishing including: knowledge/education, emotional health, physical health, physical safety, financial security, social connectedness, and spiritual practices. The interpretation of scores and correlations was based on established classifications. Findings indicated that education, family, and community support were highly valued among participants. Among the respondents, eighty-five percent reported awareness of their right to education, and 79% expressed satisfaction with their access to primary or secondary education. Most respondents felt good about their emotional health (84%). Participants felt confidence in their decision-making abilities. Though the majority of respondents recognized the importance of a balanced diet to their own physical health, less than half reported being able to make decisions about what they eat. Most respondents reported feeling safe at home, with no significant differences by gender or region. Spiritual connectedness was also strong. Many reported that spirituality brought a sense of purpose, gratitude to their lives, and inner peace and harmony. Strong spiritual values within the communities were identified as important to their wellbeing; approximately seventy-seven percent expressed comfort in speaking to their faith leaders about their problems. Financial insecurity emerged as a significant obstacle (60%), however, eighty-four (84%) expressed confidence in finding work they liked, and most (88%) believed their future work would support them financially. Findings represent a first step in gaining a cross-cultural perspective of adolescent views on human flourishing. Findings may have implications for policymakers and decision-makers seeking to understand human flourishing among adolescents, and strategies to enable their flourishing.

## 1. Introduction

Investments geared towards adolescents and youth flourishing brings benefits not only to today’s generation but also the others to come. Conversely, failure to invest in the upbringing of the young people erodes future quality and length of life. Therefore, there is a need to fill this narrative between healthy development and adolescent and youth flourishing. Moreover, missing from the body of knowledge on adolescent and youth flourishing are the dimensions of the adolescent and youth’s voice through social media, and youth engagement in community initiatives.

Research on adolescent and youth flourishing has primary conducted in developed countries with limited adolescent and youth-led analytical and cultural understanding of flourishing. Witten (2019) explains that few studies have explored the dimensions of adolescent and youth flourishing in diverse cross-cultural settings. There is an opportunity to expand current conceptualizations of adolescent and youth flourishing to enable them to voice their own perceptions of flourishing within their own contexts. Having a thorough understanding of the conditions that enable human flourishing contribute to promoting safety and security in people’s lives, including includes physical, mental, social, economic, cultural and spiritual dimensions (The Health Equity and Policy Lab, 2023). This research sought to bridge the evidence gaps and to build coherent conceptual understandings for policy recommendations and evidence building opportunities and strong theoretical underpinnings for future interventions.

## 2. Literature Review

The United Nations classifies ‘’youth’’ as those between the ages of 15.24 years. Close to 90 percent of the youth population live in less developed countries with the majority living in sub-Saharan Africa, which includes Uganda (Stecklov and Menashe-Oren 2019). This population faces specific forms of vulnerability.

For instance, adolescents are in a period of significant psychosocial development and physical growth, a pace exceeded only by the first 1,000 days of life (Stecklov & Menashe-Oren 2019). The adolescent brain undergoes tremendous growth and development, shaped by social, emotional, and behavioral exposures. As such, this period is a crucial time for education and exposure to new ideas since youth generally demonstrate significant openness to new ideas and demonstrate high levels of curiosity and interest. Despite the importance of this period in life, the recent Lancet Commission revealed significant gaps in data for indicators of child development and well-being, particularly during the period of adolescence and youth, limiting our understanding of protective and vulnerability factors that hinder healthy development.

Huppert and So (2013) define human flourishing as “a combination of feeling good and functioning effectively, and the experience that life is going well“. According to the Health Equity and Policy Lab (2023) concurs on the same and notes that human flourishing is the ability to live a good life. Rooted in Aristotelian ethics, it values health intrinsically and applies universally to all human lives. Human flourishing embraces our shared humanity and serves everyone’s interest. All people should have the conditions for flourishing and realizing their ability to be healthy; they can use their values, talents, and abilities in pursuit of their own goals and health. It is vital because it promotes the growth, development, and holistic well-being of individuals and populations. It serves as a moral basis for what it means to be a human being. For individuals’ abilities to develop, one needs to be equipped to analyze and shape health policy and public health (2023).

Templeton World Charity Foundation (2023) explains that for one to flourish, it means one is on a path towards physical and mental wellbeing that is holistically good, both for individuals and communities. Human flourishing begins with self-determination and agency, connected through strong social relationships; one flourishes when he/she lives with purpose, practices gratitude, forgives, and is open-mindedness. One cultivates character strength and resilience to flourish in adversity, to lead with empathy and virtue; one flourishes by maintaining a sense of learning, wonder, curiosity, and humility about the shared world and one’s place in it. The focus on flourishing is therefore concerned with analyzing how an extension of these interior traits, habits, and behaviors can affect one’s goals (Templeton World Charity Foundation, 2023).

Uganda has one of the youngest populations in the world, with 78 percent of its population under the age of 30. Many benefits come with a young population, but one of the most significant is the opportunity that they provide the country with to utilize their talents and energy to build a more productive and robust economy adolescents and youth are often exposed to abuse, exploitation, and violence (The Inclusive Green Economy Network, 2023). Yet as it stands in the current moment, young Ugandans are highly likely to be unemployed or underemployed. Northern Uganda in particular, experienced an 18-year period of sustained war and violence, including significant sexual violence of their reproductive health, and more broadly, their physical, cultural, economic, social and mental wellbeing (Woldetsadik, Can and Odiya, 2022).

The population also faces high levels of malnutrition during pregnancy, often leading to low birth weight, stunted growth, and infant as well as maternal mortality. Adolescents and youth in this context constitute an enormous potential for human and social development and agents of change (Zdunnek, 2003). Health and disempowerment among adolescents and youth are further exacerbated by a lack of trained personnel with multidisciplinary and comprehensive health knowledge. Today’s youth are tomorrow’s workers, entrepreneurs, and leaders, and by investing in their opportunities to build, they can shape a sustainable and thriving future for themselves (The Inclusive Green Economy Network, 2023). This research therefore sought to understand the determinants that affect youth flourishing in Mayuge and Mpigi Districts in terms of providing them with opportunities to shape a sustainable and thriving future.

## 3. Objective of the Study

With focus on youth, the research sought to build a shared understanding the determinants of human flourishing by exploring and establishing the relationship between adolescent flourishing in diverse cross-cultural settings, gender, social, economic and spiritual wellbeing in Mayuge and Mpigi districts, in Uganda.

### 3.1. Specific Objectives

The specific objectives are:

1. To gain the perspectives of adolescents and youth in Mayuge and Mpigi in Uganda on human flourishing and its relationships to the dimensions of human flourishing.
2. To contribute evidence through emerging literature on the interconnectedness between health seeking behaviors, health care delivery, and human flourishing among adolescents and youth in highly vulnerable socio-economic contexts.
3. To understand and develop strategies for adolescent and youth flourishing that can guide our understanding of adolescent and youth strategies for flourishing in diverse cross-cultural contexts.

## 4. Methodology

This quantitative cross-sectional study was conducted in Mpigi and Mayuge districts in Uganda between November and December 2022. The study included adolescent boys and girls aged 14-19 years. Recruitment methods involved community engagement, school staff, youth networks, healthcare facilities, and village health teams. A total of 333 adolescents were targeted for participation, with approximately 167 from each study site. Interviews were conducted and they involved 55 students who were selected from six secondary schools. The sample size was drawn from an estimated total of about 2,500 students in the sampling frame from the communities. Convenience sampling was employed, among students who expressed willingness to participate and had parental consent to be included in the study.

Data collection was done electronically using ODK electronic data capture platform with tablets running V10.1.1 operating system. Data collectors were trained on the data collection procedures, how to use tablets for data collection, data transmission and troubleshooting in case of any errors or technical challenges during data collection. The data was subjected to data quality control processes and preparation for analysis (data cleaning). Data cleaning and analysis was done in STATA v17.

Descriptive statistics were computed to summarize variables, including frequency counts and proportions for categorical variables and summary statistics (mean, standard deviation for normally distributed variables or median and interquartile range for non-normally distributed variables) for continuous variables. Disaggregated data was analyzed using independent sample T-tests and χ2 tests (or Fisher’s exact test for variables with a cell count of <5). Composite scores were generated for each theme by averaging individual scores, allowing for comparison on the same scale. Cronbach’s alpha coefficient assessed internal consistency, and pairwise Karl Pearson correlation coefficient examined relationships between themes. The interpretation of scores and correlations was based on established classifications.

## Ethics Statement

The research was approved by the University of Saskatchewan Behavioural Research Ethics Board (Beh-REB) under Application ID no 1996. The University of Saskatchewan Behavioural Research Ethics Board (Beh-REB) is constituted and operates in accordance with the current version of the Tri-Council Policy Statement: Ethical Conduct for Research Involving Humans - TCPS 2 (2018). The University of Saskatchewan Beh-REB has reviewed the above-named project. The proposal was found to be acceptable on ethical grounds. The principal investigator has the responsibility for any other administrative or regulatory approvals that may pertain to this project, and for ensuring that the authorized project is carried out according to the conditions outlined in the current approved protocol. This Certificate of Approval is valid for the above time period provided there is no change in experimental protocol or consent process or documents. Furthermore, formal verbal consent was obtained for adult human participants while written informed consent was obtained from the parent/guardian of each participant under 18 years of age during the study investigation.

## 5. Findings

This section presents the research findings.

### Respondents’ Socio-Demographic Characteristics

Table 1 presents socio-demographic characteristics of participants who took part in the survey. A total of 299 respondents took part in the survey (a response rate of 90%). Out of these, 46% were male 52% female with only about 2% cohabiting with someone with only about 2% cohabiting with someone. The average number of household members where the respondents lived was 7.5 individuals. Most of the participants (53%) lived with both parents, 39% with one parent and 6% lived with non-family members. Only about a third (32%) of the respondents were involved in income generating activities with the average monthly income per household being reported as 11 United States Dollar (USD).

**Table 1:** Participant’s Socio-demographic Characteristics.

|  | All<br>n (%) | Mayuge<br>n (%) | Mpigi<br>n (%) |
| --- | --- | --- | --- |
| Gender |  |  |  |
| *Boys* | 136 (45.5) | 70 (46.7) | 66 (44.3) |
| *Girls* | 163 (54.5) | 80 (53.3) | 83 (55.7) |
| Marital Status |  |  |  |
| *Single* | 291 (97.3) | 142 (94.7) | 149 (100) |
| *Cohabiting* | 6 (2.0) | 6 (4.0) | 0 |
| *Ever Married* | 2 (0.7) | 2 | 0 |
| Household size (mean (sd)) | 7.5 (3.3) | 8.0 (3.6) | 7.0 (2.9) |
| Household size (median (IQR)) | 7 (6-10) | 7 (6-8) | 7 (5-8) |
| Current living situation |  |  |  |
| *Living with one parent* | 117 (39.1) | 44 (29.3) | 73 (49.0) |
| *Living with both parents* | 159 (53.2) | 87 (57) | 72 (48.3) |
| *Living with non-family members* | 19 (6.4) | 18 (12) | 1 (0.7) |
| *Other living arrangements* | 5 (1.7) | 2 (1/3) | 3 (2.0) |
| Involved in income-generating activities | 97 (32.4) | 46 (30.7) | 51 (34.2) |
| Average income in USD (Median (IQR)) | 11 (4-19) | 11 (7-19) | 8 (4-19) |
| Schooling level |  |  |  |
| *Primary/elementary* | 17 (5.7) | 17 (11.3) | 0 |
| *Secondary/high school* | 282 (94.3) | 133 (88.7) | 149 (100.0) |
| Attending a place of worship |  |  |  |
| *Never* | 8 (2.7) | 6 (4.0) | 2 (1.3) |
| *Less than once a year* | 15 (5.0) | 7 (4.7) | 8 (5.4) |
| *Several times a year* | 64 (21.4) | 33 (22.0) | 31 (20.8) |
| *Once a month* | 30 (10.0) | 16 (10.7) | 14 (9.4) |
| *About weekly* | 182 (60.9) | 88 (58.7) | 94 (63.1) |
| Volunteering |  |  |  |
| *Never* | 94 (31.4) | 47 (31.3) | 47 (31.5) |
| *Less than once a year* | 30 (10.0) | 12 (8.0) | 18 (12.1) |
| *Several times a year* | 88 (29.4) | 55 (36.7) | 33 (22.2) |
| *Once a month* | 42 (14.1) | 13 (8.7) | 29 (19.5) |
| *About weekly* | 39 (13.0) | 19 (12.7) | 20 (13.4) |
| Average number of close friends (median (IQR)) | 3 (2-5) | 2 (1-3) | 2 (1-3) |
| Religion |  |  |  |
| *Christian* | 168 (56.2) | 84 (56.0) | 84 (56.4) |
| *Muslim* | 130 (43.5) | 65 (43.3) | 65 (43.6) |
| *Other* | 1 (0.3) | 1 (0.7) | 0 (0.0) |

Majority of the respondents (94%) were in secondary/ high school level, 56% were Christians while 44% Muslims. Most of the respondents (61%) indicated that they attended a place of worship at least once a week followed by those who attended several times a year at 21%. Additionally, slightly less than a third (29%) of respondents volunteered to do an activity several times a year, 14% once a month and 13% about weekly. Respondents also reported to have (median number of 3) (IQR: 2-5) close friends-3.

Most of the respondents interviewed were single boys and girls in secondary school living with one or both parents. The sample comprised of both Christians and Muslims who attended a place of worship at least once a week. Most of the respondents were not involved in any income generating activities. Moreover, the education level of the respondents validates the data with the majority having secondary/high school level of education; therefore, they were able to understand the questionnaire and answer accordingly.

Respondents were asked to rate their level of agreement to a set of questions under eight identified areas. These included: knowledge/education, emotional health, physical health, physical safety, financial security, social connectedness, spiritual purpose and equality measures. Findings under each of the eight themes are presented below with a subsequent examination of the relationships among the themes.

### Knowledge/Education

**Table 2** presents the findings of the eight questions used to measure knowledge/education theme. The results were also compared by region and gender. Overall, a total of 91% of respondents agreed that they attended school every day (93% in Mayuge while 89% in Mpigi, p<0.01). Almost all respondents (97%) believed that education will give them a bright future, and this was similar both by gender (p=0.21) and by region (p=0.22). Only 2.7% were not sure about the benefits of education. Another item assessed was whether their family encouraged education regardless of what gender they were. Majority (93%) agreed with just slightly more female (96%) than male (90%), p=0.07. Thus, the difference by region was minimal.

**Table 2:**
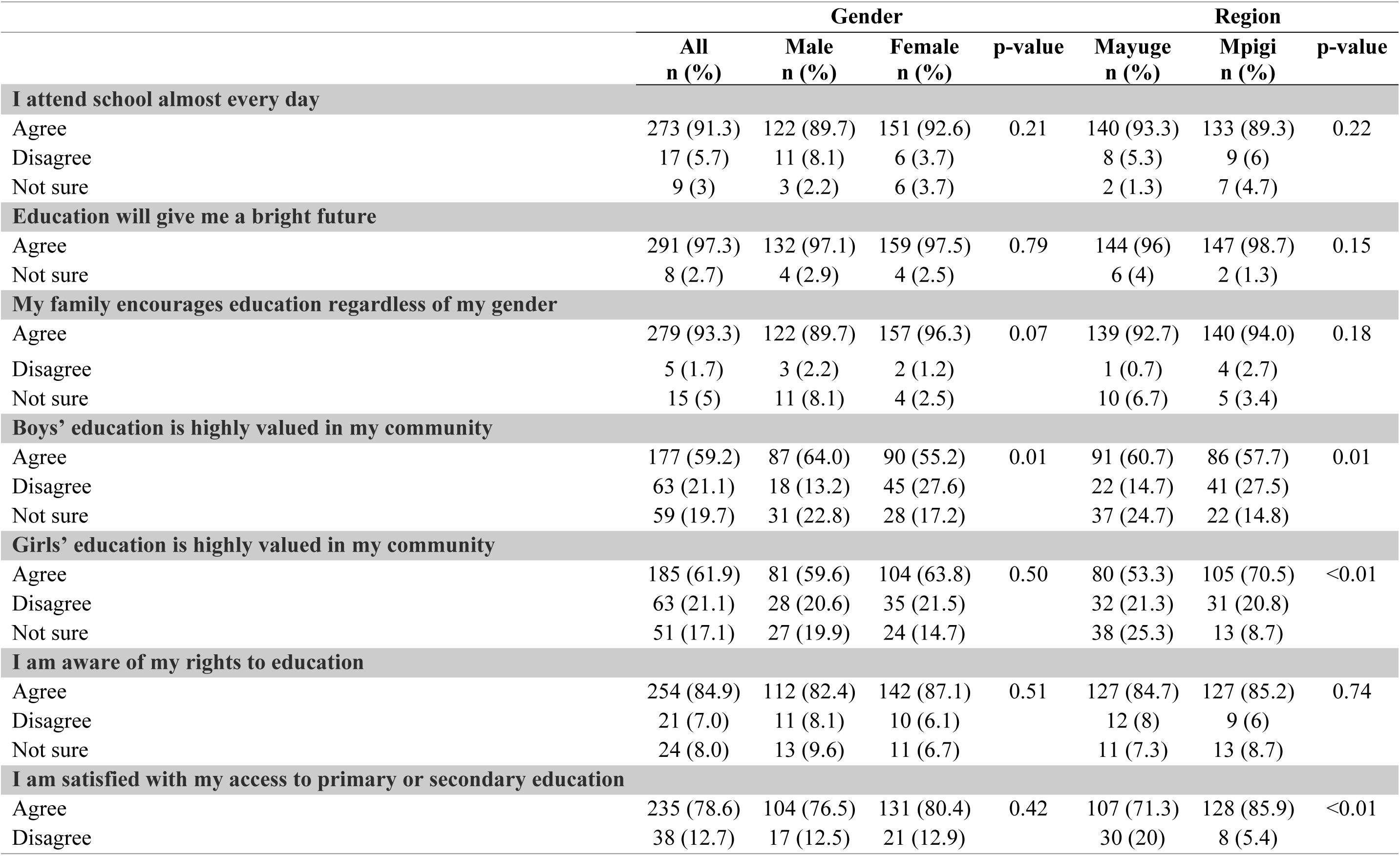
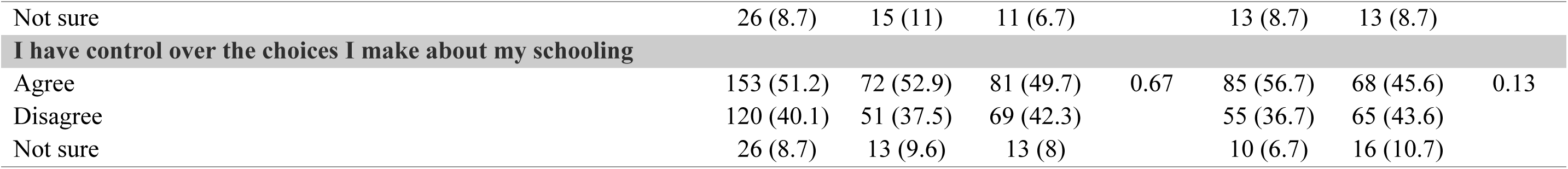
Knowledge/education thematic area scores disaggregated by gender and region.

On whether young men’s education was highly valued in the community, close to 60% of all respondents with more males (64%) compared to young women’s (55%) agreed about this (p=0.01). Slightly over a quarter (27%) of the young women compared to 13% of the young men who disagreed, and a relatively higher proportion of young men (23%) compared to young women (17%) were not sure. Regionally, there was no significant difference in the proportion of those who agreed (Mayuge=61%, Mpigi 58%); however, a higher proportion of those from Mpigi (28%) compared to those in Mayuge (15%) disagreed with close to a quarter in Mayuge being unsure about the privilege of male education over female schooling(p=0.01). A total proportion of 62% of respondents agreed that girls’ education was highly valued in the community and no significant difference by gender (p=0.50) was noted. Nonetheless, the proportion of those who agreed that girls’ education was valued in the community was higher in Mpigi (71%) compared to Mayuge (53%) where again a quarter of the respondents (25%) were not sure. These findings imply that the communities in both Mayuge and Mpigi did acknowledge the importance of education, especially girl education as a key factor to a better future. These results indicate a cultural shift from past years whereby girls’ education was not valued.

The importance of education and especially girls’ education is also confirmed by the same majority who reported the need to recognize the right to education for all. According the findings, respondents’ knowledge about their rights to education was also assessed with 85% agreeing that they were aware, 8% were unsure and a considerable 7% reported that they were not aware. There were no major differences by gender and by region as well. Additionally, respondents were asked to score whether they were satisfied with their access to primary or secondary education. A total of 79% agreed with no significant differences by gender (p=0.42), but had higher proportion of respondents from Mpigi (86%) compared to Mayuge (71), p<0.01. On control over choices made about school, about half of the respondents (51%) reported that they agreed and there were no significant differences by gender (p=0.67) and region (p=0.13).

From the results, most respondents reported attending school every day, driven by the belief that education is a path to success. The respondents opined that girls’ and boys’ education was highly valued and that the two communities under study value education irrespective of the gender; therefore, almost all respondents’ families encouraged education. Results also show that most of the respondents are aware of their right to education and showed satisfaction with access to school. The remaining number of respondents who did not value girls’ education is an indication of the continuing struggle for gender equity. However, over half of the respondents noted that they had control over choices made about school while the rest indicated that they had no decision-making power over their education.

### Emotional Health

Emotional health is an important thematic area for adolescent flourishing. It is about having the skills and resources to manage the ups and downs of every day-to-day life. Having good emotional health is a fundamental aspect of fostering resilience, self-awareness, and overall contentment. To measure emotional health among the youth, the study considered the following indicators: feeling good about life, life purpose, being confident about one’s decisions, being a good person, hopefulness, doing things well, smiling, sadness, where to turn to for in case of problems, usually expecting to have a good day, being left out of group activities, unwanted sexual advances and emotional health to be central to a bright future. **Table 3** presents the findings on emotional health amongst youth.

**Table 3:**
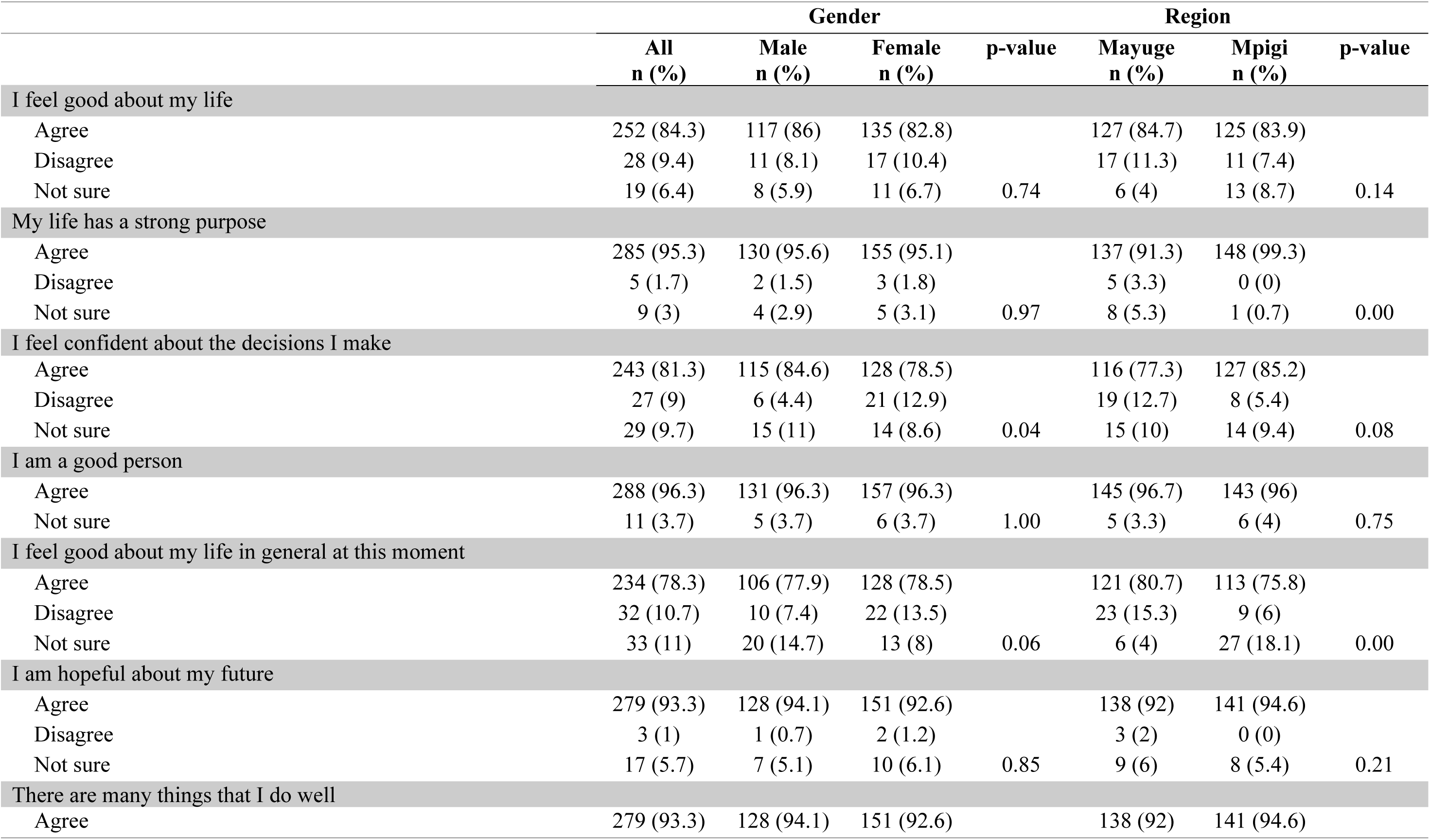
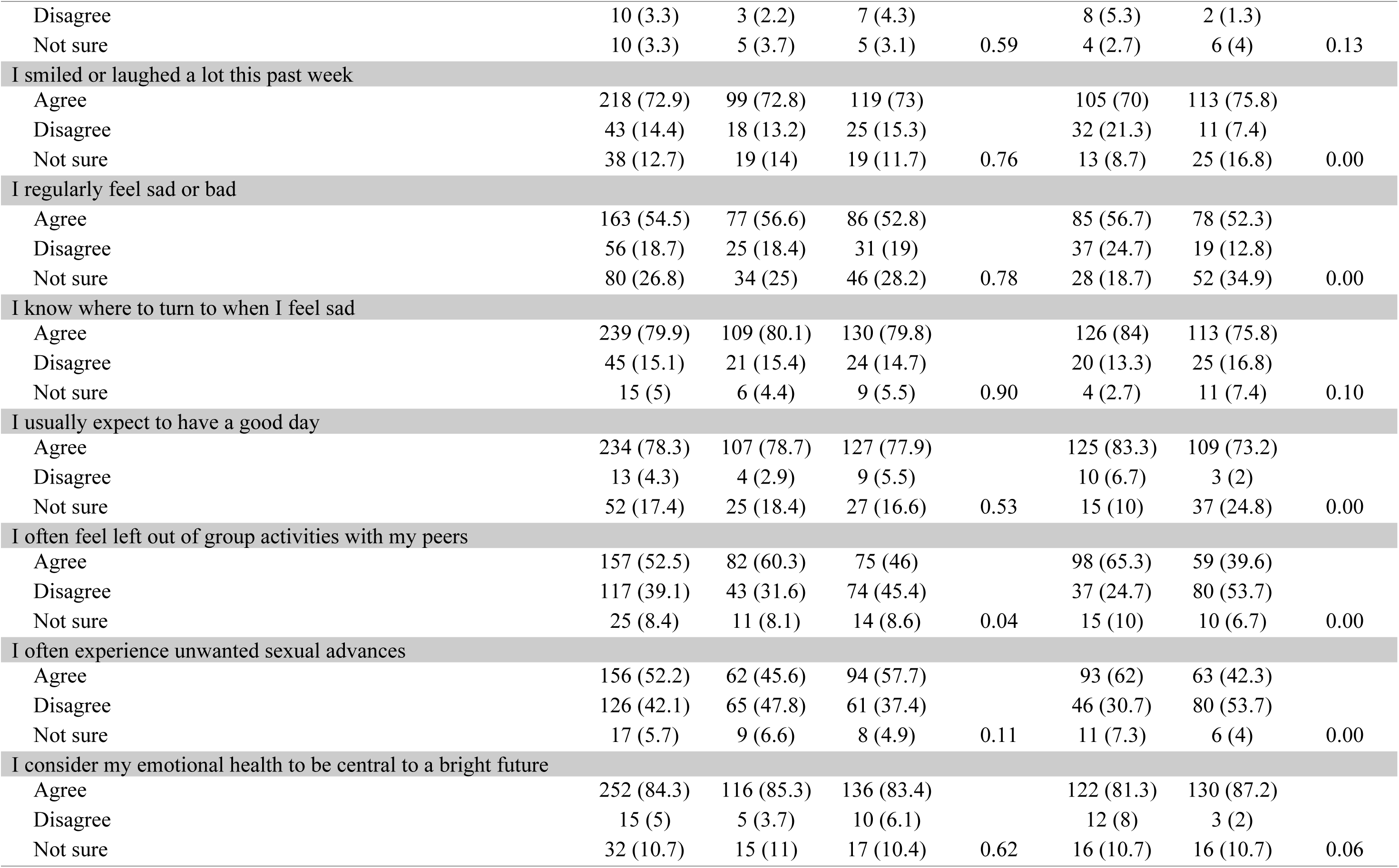
Theme; Emotional health scores disaggregated by gender and region.

Most of the respondents who took part in the survey (84%) agreed that they felt good about their life with no significant difference by gender (p=0.74) and region (p=0.14). A total of 95% agreed that their life had a strong purpose with no difference by gender (p=0.97) but the proportion was higher in Mpigi (99%) compared to Mayuge (91%), p<0.01.

Over three quarters of the respondents (81%) agreed that they felt confident about the decisions they made for themselves with Mpigi (85%) recording a higher proportion than Mayuge (77%) and higher proportion among male (85%) compared to female (79%). When asked to rate their character, most of the respondents (96%) perceived themselves as good individuals, with no difference by gender (p=1.00) and region (p=0.75). As reported by respondents, 78% indicated that they felt good about their life in general at the time of the survey. No major difference between male and female among those who reported that they felt good but slightly more female (14%) than male (7%) reported not feeling good about themselves, with 15% among the male reporting that they were not sure. Comparing regions, 80% of respondents in Mayuge agreed that they felt good about their life in general now compared to 76% from Mpigi. Notably, 15% in Mayuge reported that they were not feeling good compared to 6% in Mpigi where a relatively higher proportion (18%) were indifferent about this statement.

In terms of hope for the future, 93% (n=279) of the total respondents agreed that they were indeed hopeful about their future with no difference by gender (p=0.85) and region (p=0.21). Across the board, most of the respondents (93%) agreed that there are many things they do well with and there was no difference by gender (p=0.59) and region (p=0.13). Also, most respondents (73%) agreed with the statement that they had smiled or laughed a lot in the past week with no difference by gender (p=0.76) but Mpigi had a relatively higher proportion (76%) than Mayuge (70%). Notably, 21% in Mayuge compared to only 7% in Mpigi reported not to have smiled or laughed a lot in the past week.

Out of all respondents, slightly over half (55%) agreed that they regularly felt sad or bad with a considerable proportion (27%) reporting that they were unsure. There was no difference by gender but the proportion of those who were unsure in Mpigi (35%) was almost twice as much of those from Mayuge (19%), p<0.001. The majority (80%) agreed that they had a person to turn to when they felt bad or sad. Slightly over three-quarters (75.8%) of all respondents usually expected to have a good day with more respondents from Mayuge (83%) compared to Mpigi (73) agreeing to this statement (p<0.001).

On whether they often felt left out of group activities with peers, half of all respondents (52%) agreed, a majority being boys (60%) compared to girls (46%), p=0.04 and majority in Mayuge (65%) compared to Mpigi (40%). When asked to score whether they often experienced unwanted sexual advances, 53% of the respondents agreed, the majority being girls (58%) compared to boys (46%), and this proportion was higher in Mayuge (62%) compared to Mpigi (42%). Lastly, on this theme, respondents were asked whether they considered their emotional health to be central to a bright future. A total of 84% agreed with no difference by gender (p=0.62) but was higher in Mpigi (87%) compared to Mayuge (81%).

A majority of the respondents interviewed indicated that they were happy about their lives and what they did in life. Most of the respondents expressed confidence in the decisions they made and felt that they were good people. In addition, a bigger percentage of the respondents pointed out that they had a strong purpose in life and were hopeful in life with a larger percentage of the respondents considering their emotional health to be central to a bright future. The data showed that most of the respondents expected to have a good day. However, slightly over half of the respondents indicated that they regularly felt sad but then most of them had a place to turn to when they felt this way. Findings also indicated that half of the respondents were left out of peer group activities, with the larger percentage being boys. In addition, slightly over half of the respondents, mostly girls, experienced unwanted sexual advances.

These findings imply that most of the youth expressed happy emotions and reported having the skills and resources to manage the ups and downs of every day-to-day life, which enabled them to foster resilience, self-awareness, and overall contentment in their life. Furthermore, the correlational analysis of the indicators confirmed a positive relationship between the indicators of emotional health. This means the higher the indicators, the stronger the emotional health among the youth.

### Physical Health

Physical health is another key element to flourishing in life, particularly for adolescents who are still growing and maturing. There is a very strong positive relationship between the two variables (physical health and flourishing in life). Indeed, one must be in good health to be able to achieve one’s set goals. Physical health can be defined as the normal functioning of the body, representing one dimension of total well-being. It is about body physical wellbeing and ability to perform day-to-day activities. As for this study, physical health was assessed using a set of 8 items, namely, balanced diet, making one’s own decision about what to eat, access to clean water, practice of basic sanitation, being able to solve one’s health problems, sharing one’s concerns, awareness of safe sexual practices and believe in safe sexual practices. The findings are presented in Table 4.

**Table 4:**
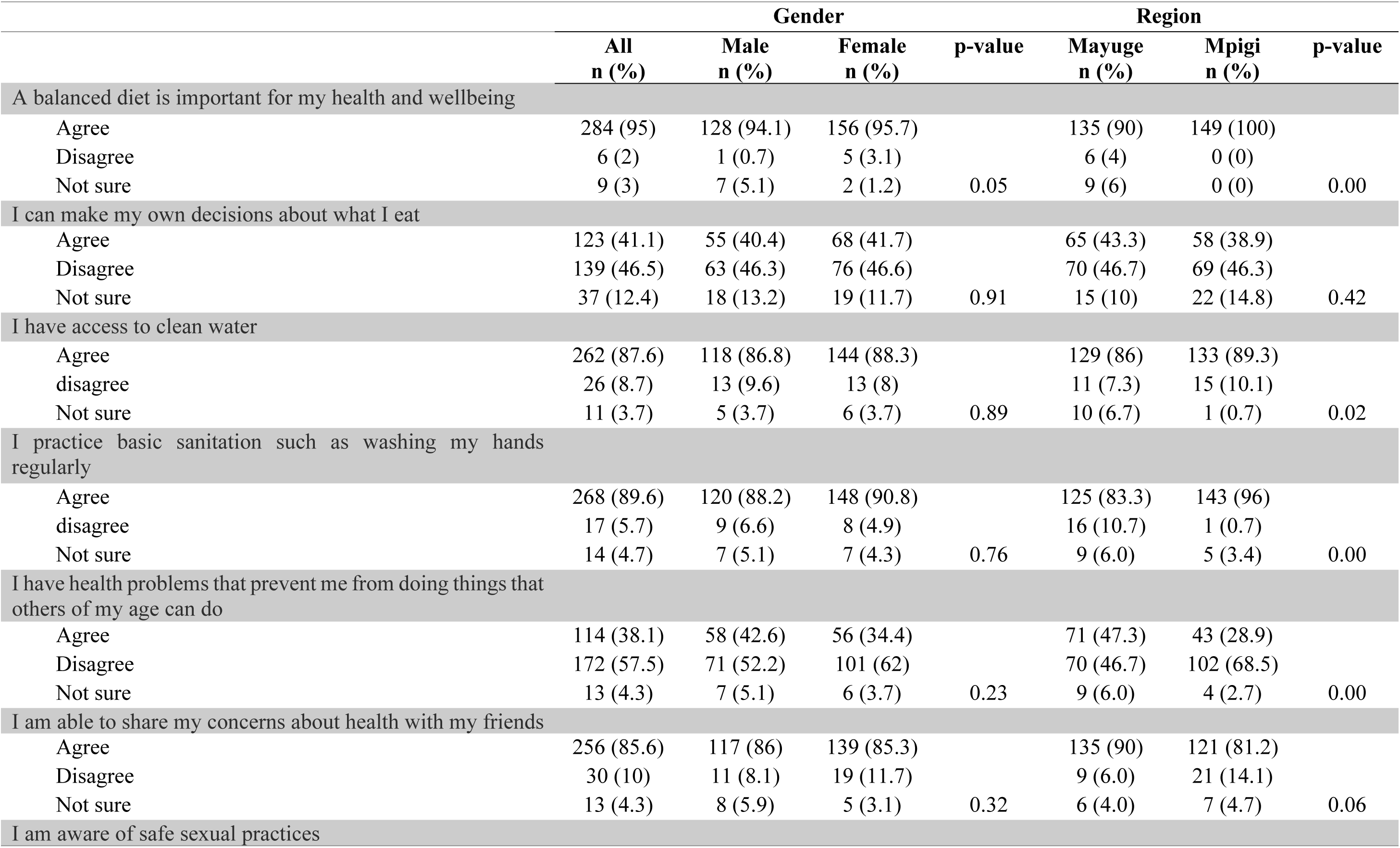
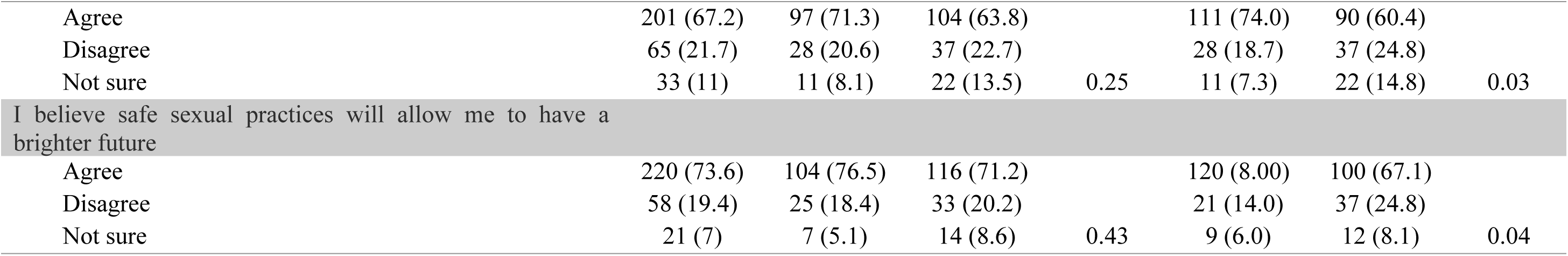
Theme Physical Health scores disaggregated by gender and region.

A total of 95% of the respondents reported that a balanced diet was important for their health and wellbeing. Regarding decisions about what they ate, there was even distribution in the proportion of those who agreed (41%) and those who disagreed (47%) with 12% being not sure. There was no difference by gender and region. A total of 88% reported that they had access to clean water, and the proportion was slightly higher in Mpigi (89%) compared to Mayuge (86%). A total of 90% of respondents reported that they practiced basic sanitation such as washing hands regularly with this proportion again being higher in Mpigi (96%) compared to Mayuge (83%). Out of all respondents, 38% reported that they had a health problem that prevented them from doing things that other people of the same age could do, this proportion being highest in Mayuge (47%) than Mpigi (29%).

A total of 86% of respondents agreed that they were able to share their concerns about their health with their friends. There was no significant difference by gender (p=0.32) but there was a marginal difference by region (Mayuge 90% vs Mpigi 81%, p=0.06). A total of 67% of the respondents were aware of safe sexual practices and 74% believed that safe sexual practices would allow them to have a brighter future. The proportion of those who believed in the importance of safe sexual practices was higher in Mayuge (80%) compared to Mpigi (67%).

Almost all respondents agreed that a balanced diet was important for their health and well-being. However, only about half of the respondents reported being able to decide on what they eat. Concerning sanitation and hygiene, the majority of the respondents reported to have access to clean water with most of them practicing basic hygiene. Some participants noted that they had health problems that prevented them from performing tasks compared to their counterparts. However, most of them were able to share their health concerns with their friends.

The respondents were also aware of safe sexual practices with a majority believing that safe sexual practices would allow them to have a brighter future. Furthermore, the analysis also revealed that while the youth are aware of the importance of a balanced diet, they are not able to decide what to eat. Furthermore, while they were aware and believed in the importance of safe sex practices, the study could not establish whether participants from the two regions experienced good physical health and refrained from risky sexual behaviors.

### Physical Safety

Physical safety is experienced when the likelihood of harm or injury is minimal or non-existent. The concept of physical safety is strongly connected to psychological and emotional safety. The study considered the following indicators to measure physical safety of the respondents. Notably: safety at home, safety at school, safety in the community, confidence in my ability to refuse unwanted sexual advances, knowledge on how to find help when unsafe, understanding one’s rights to sexual and reproductive health, feeling unsafe because of one’s gender, and importance of safety to one’s well-being.

Table 5 presents the results. Respondents were asked to state whether they felt safe in their homes, with 84% of them agreeing, but there was no significant difference by gender (p=0.63) or region (p=0.21). A high proportion of 88% felt safe at school, but this was varied by gender with more young women (10%) compared to young men (4%) reporting feeling unsafe. The proportion of those who felt unsafe in school was higher in Mayuge (14%) with 81% feeling safe compared to Mpigi’s 1% (unsafe) and 95% (safe). When asked about the level of safety they felt when walking in their communities, only slightly over half (56%) agreed that they did feel safe, with marked differences by gender (70% male vs 45% female, p<0.01) and region (61% Mayuge vs 52% Mpigi, p=0.01).

**Table 5:**
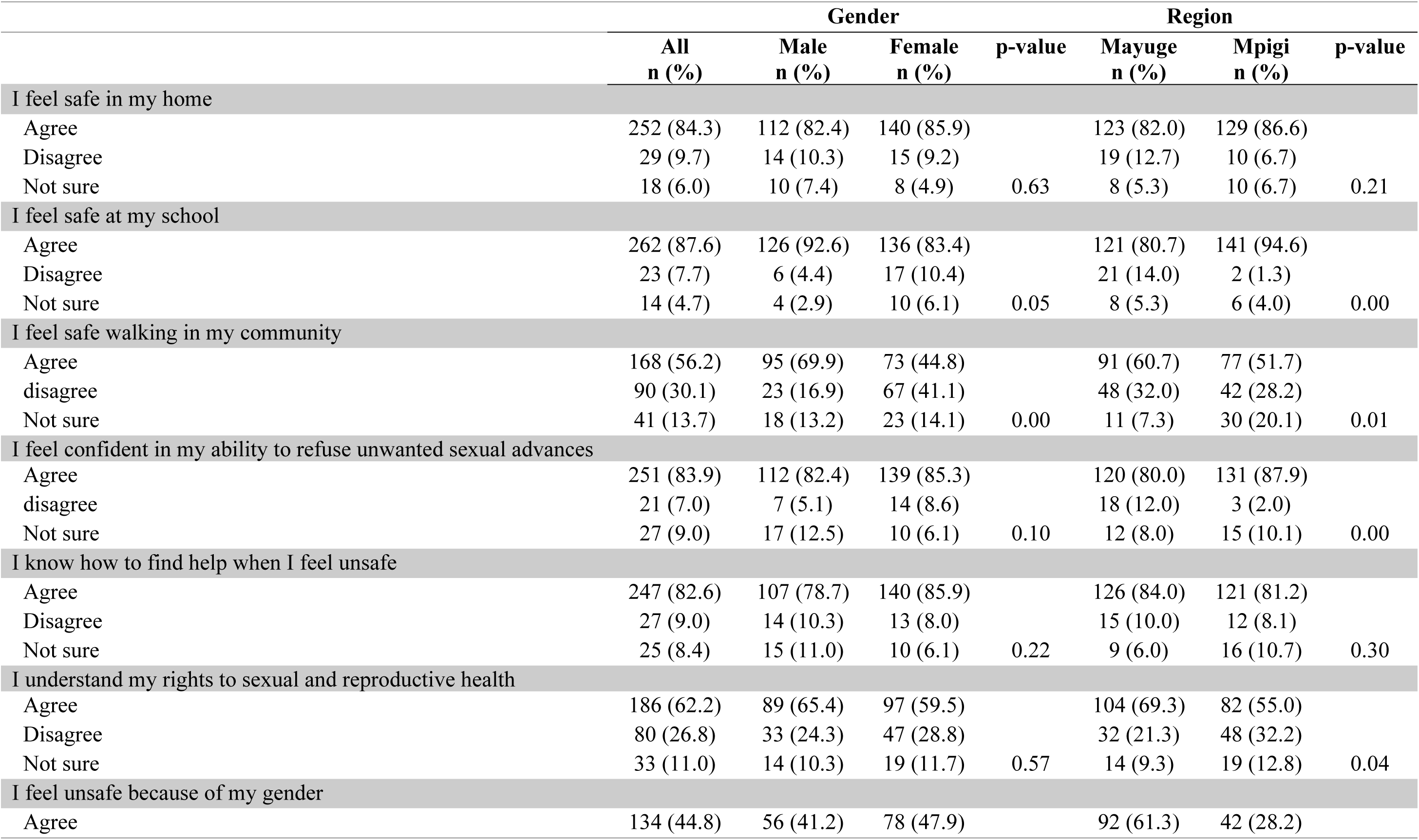
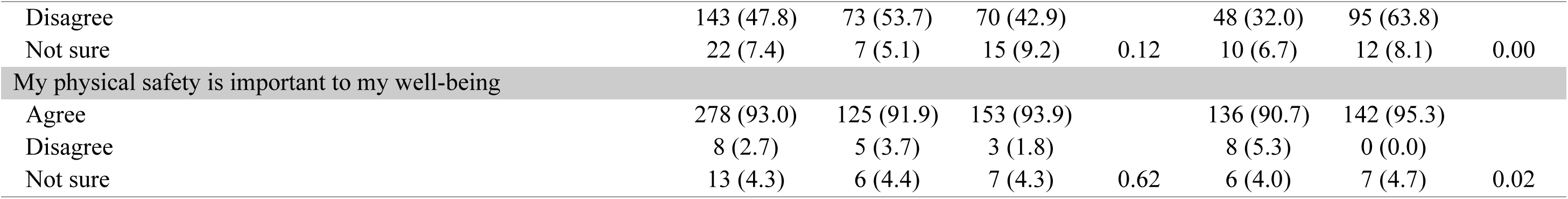
Theme Physical Safety scores disaggregated by gender and region.

This implies that more young women (41%) than young men (17%) and more from Mayuge (32%) than Mpigi (28%) felt unsafe walking in their community. Additionally, out of all respondents who took part in the survey, 84% agreed that they felt confident in their ability to refuse unwanted sexual advances, while 83% knew how to find help when they felt unsafe. Only 62% reported that they understood their rights to sexual and reproductive health with this proportion being different by region (69% in Mayuge vs only 55% in Mpigi, p=0.04. Slightly less than half (45%) of the respondents reported that they felt unsafe because of their gender; however, the difference was not so significant.

Nonetheless, this varied by region with more feeling unsafe because of their gender in Mayuge (61%) compared to Mpigi (28%). The survey sought to also establish how respondents would score their perception of the importance of their physical safety to their well-being. A high proportion (93%) agreed that their physical safety indeed contributed to their well-being with no difference by gender (p=0.62) but slightly higher in Mpigi (95%) compared to Mayuge (91%), p=0.02.

Generally, the results show that respondents felt safe both at home and school. However, a higher percentage mentioned that they felt safe at school. Slightly over half of the respondents indicated feeling safe while walking in the community with women expressing safety concerns while walking in the community. Additionally, under half of the participants, mostly from Mayuge reported feeling unsafe because of their gender. Participants felt confident in their ability to refuse unwanted sexual advances and mentioned that they knew how to find help when they felt unsafe. Further, the study respondents pointed out that physical safety does contribute to their well-being. In addition, they reported that they understood their rights to sexual and reproductive health.

### Financial Security

Financial security is defined in a variety of ways by different people. However, it generally refers to the peace of mind felt when one is not worrying about one’s finances. Often, this means having an adequate income to comfortably cover expenses, being debt-free, and having savings for emergencies. Despite the varied understandings of the term, literature confirms a strong positive relationship between financial security and flourishing in life (VanderWeele, 2017). The study considered the following indicators to measure the status of financial security among the youth of Mayuge and Mpigi, in Uganda: meeting basic needs every day, meeting monthly living expenses, ability to find work that one likes doing, financial support from future work, ability to support my family financially in the future, not having enough money being a stress, being “poor” in comparison with others, and being “rich” in comparison with others.

Table 6 presents results from all the eight items used to measure financial security. Out of all respondents interviewed, 55% agreed that their basic needs were met every day, this proportion being higher in Mpigi (66%) compared to Mayuge (44%), p<0.01. Slightly over half 54% of respondents reported that they were worried that their household could not meet monthly living expenses, but 84% expressed confidence that they would be able to find work that they liked doing. An analysis by region showed that the proportion of those who expressed confidence in finding work was higher in Mpigi (91%) compared to Mayuge (76%). The majority of respondents (88%) agreed that they were confident that their future work would support them financially, with this proportion again being higher in Mpigi (97%) compared to Mayuge (79%). Similarly, 87% of the respondents reported having confidence that they would be able to support their families financially in the future - this proportion being higher in Mpigi (95%) compared to Mayuge (79%). Not having enough money was a major stress for 60% of the respondents (67% in Mayuge vs 52% in Mpigi, p=0.04).

**Table 6:**
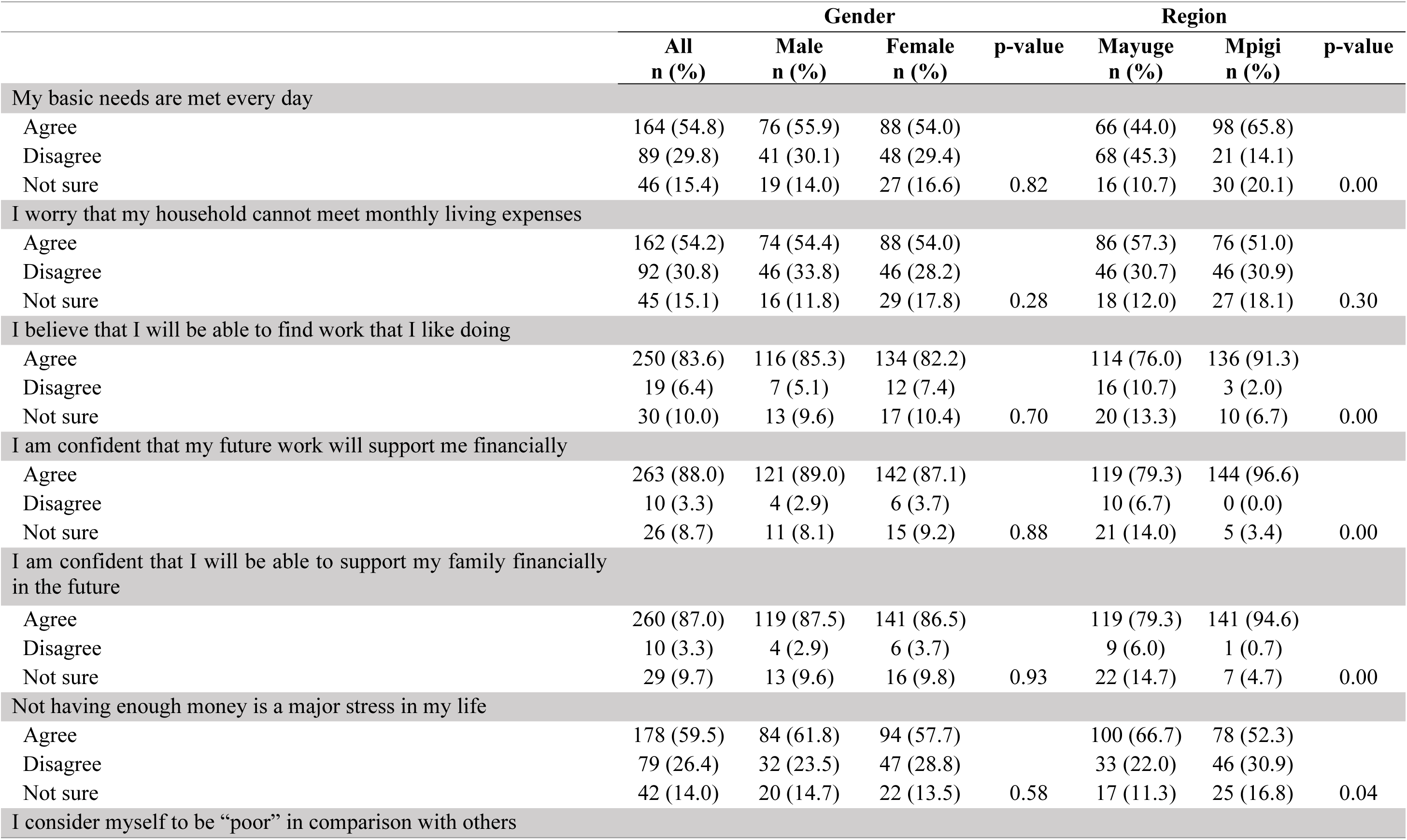
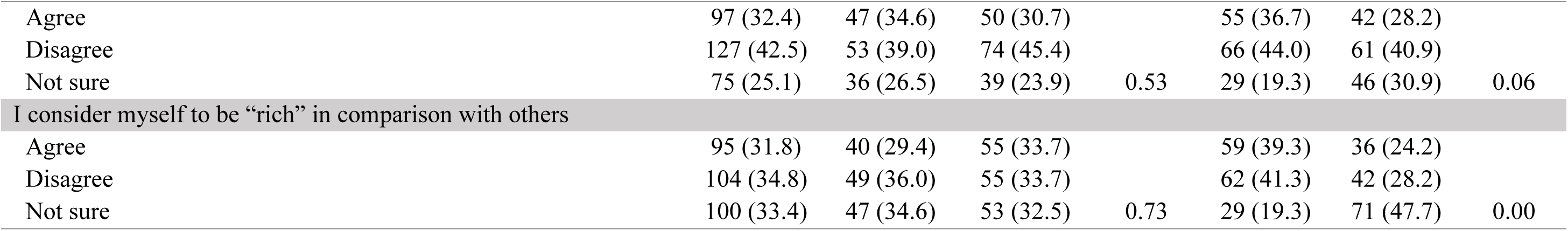
Theme Financial Security scores disaggregated by gender and region.

About a third (32.4%) of respondents considered themselves “poor” in comparison to others with this proportion being marginally higher in Mayuge (37%) compared to Mpigi (28%), p=0.06. Another a third (32%) considered themselves “rich” in comparison to others with this proportion again being slightly higher in Mayuge (39%) compared to Mpigi (24%), p<0.01. Notably though, a high proportion of respondents from Mpigi were unsure on how to respond to the question on whether they considered themselves “rich” in comparison to others.

The results show that more than half of the respondents were able to meet their basic needs; however, most of them expressed concern that their households could not meet their monthly living expenses. Respondents were confident about getting work that they like, which would support them and their families financially. Additionally, the respondents expressed mixed reactions when asked whether they considered themselves poor or rich in comparison to others with the majority of them indicating that lack of money was a major stress for them. Furthermore, the analysis in Table 7 also confirmed a strong positive relationship between financial security and flourishing in life. All the variables revealed a positive correlation with financial security, meaning that the more these variables become stronger in the life of youth, the better their flourishing and well-being. In other words, a youth who is unable to afford basic needs and meet his/her monthly living expenses because of lack of money will likely not flourish and be well in life.

**Table 7:**
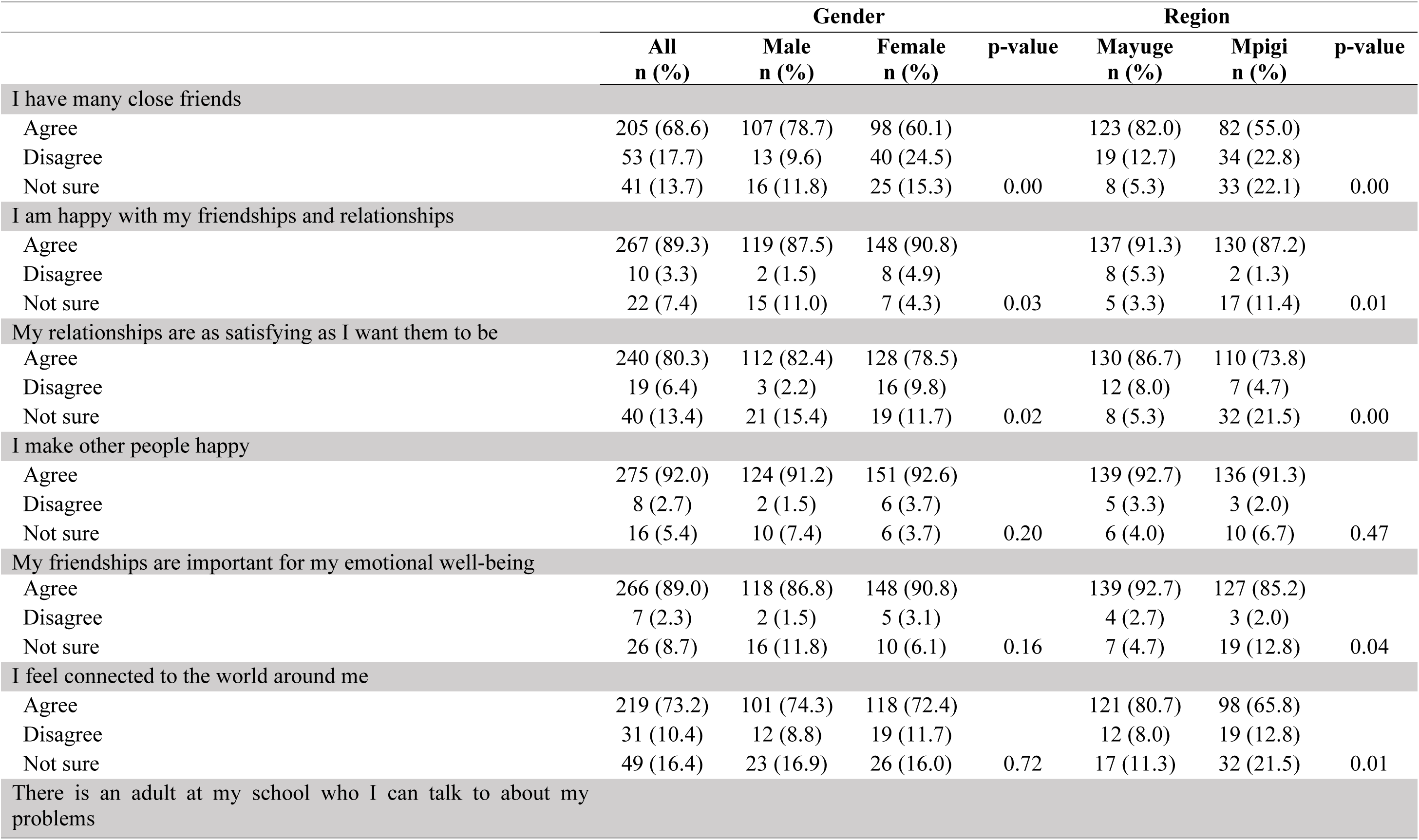
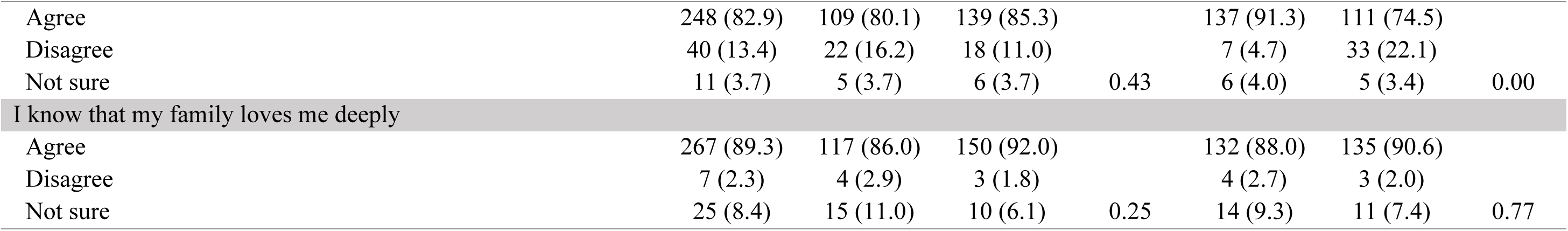
Theme Social Connectedness scores disaggregated by gender and region.

### Social Connectedness

For this research, social connectedness refers to one’s ability to have relationships with others which may consist of immediate/extended family, friends, and the community at large. It is about the relationships people have with each other and their engagements with the broader community. Social connection is an integral component of health and well-being. It can lower anxiety and depression, help us regulate emotions, lead to higher self-esteem and empathy, and improve immune systems. By neglecting our need to connect, we put our health at risk.

Social connectedness was examined using a set of eight indicators. Namely: having many close friends, being happy about friendships and relationships, being satisfied with one’s relationships, making other people happy, acknowledging the importance of one’s friendships for one’s emotional well-being, feeling connected to the world around, having an adult at my school who I can talk to about my problems, and feeling deeply loved by one’s family. The findings are presented in **Table 7**.

A total of 69% of respondents agreed that they had many close friends compared to a combined proportion of 18% respondents who disagreed. There was significant difference by region with those in Mayuge (82%) rating higher than those in Mpigi (55%), p<0.01. Again, the proportion was higher among the male (79%) compared to female (60%), p<0.01. Out of all the respondents who took part in the survey, 89% agreed that they were satisfied with their friendships and relationships and 80% further agreed that their relationships were as satisfying as they wanted them to be (82% male vs 79% female, p=0.02). This proportion was higher in Mayuge (87%) than Mpigi (74%). The majority of respondents (92%) opined that they made other people happy and a further 89% reported that their friendships were important for their emotional well-being. There were no significant differences by gender and region in these two items.

When asked whether they felt connected to the world around them, 73% of all respondents confirmed having friendships, more in Mayuge (81%) compared to Mpigi (66%). A total of 83% of all respondents reported that there was an adult in school who they would talk to about their problems, more so in Mayuge (91%) compared to Mpigi (75%). Lastly on this theme, 89% of all respondents reported that they knew their family loved them deeply.

Most respondents reported having many close friends, this was most evident among male respondents with a majority saying that they were satisfied with their friendships and relationships and that they also made other people happy. Still, a higher proportion of the youth interviewed pointed out that their friendships were important for their emotional well-being. In addition, most respondents mentioned that they felt connected to the world and that there was an adult in their school who they would talk to about their problems. Lastly, most respondents indicated that they knew that their families loved them deeply. The analysis of these findings therefore confirms the element of social connectedness among the youth in Mayuge and Mpigi contributing to their flourishing. As a result of the social connectedness, the youth can lower anxiety and depression, help regulate their emotions, have higher self-esteem and empathy, and improve their immune systems.

### Spiritual Purpose

Spiritual purpose is not related to anything material. It does not involve issues such as one’s career or where one’s live. On the contrary, it is about establishing a set of principles, values and beliefs that give one’s life meaning and then using them to guide the decisions and actions that one makes. Spiritual purpose helps one to become one’s best self as one moves through life. One can find higher purpose in life through many different pathways, including meditation and prayer, personal reflection and practicing spiritual wellness.

The study sought to establish whether the youth in Mayuge and Mpigi made use of spiritual purpose to establish a set of principles, values, and beliefs to give them a sense of meaning in their lives. To do so, seven indicators were considered, namely, importance of spiritual purpose as part of daily life, sense of purpose to life, feeling grateful for life, deep inner peace or harmony, sharing spiritual values with the community, speaking to a faith leader about one’s problems, and connection to all life.

Results from the seven items used to measure spiritual purpose theme are presented in **Table 8**. Out of all respondents, 99% agreed that spirituality was an important part of their lives with this being similar across both genders (p=0.30) and across both regions (p=0.13). For most respondents (94%), spirituality helped to bring a sense of purpose to their life (Mayuge, 89% vs Mpigi, 99%, p<0.01) and 82% reported that they felt grateful for their lives i.e being able to wake up every day (Mayuge, 80% vs Mpigi, 85%, p=0.01). When asked to score their sense of inner peace and harmony, 75% of the respondents were on the affirmative with 12% among the female compared to only 4% among the male reporting that they did not feel deep inner peace or harmony (p=0.02. In total, 78% of respondents agreed that their spiritual values are shared by the communities they dwell in with more respondents being from Mpigi (81%) compared to Mayuge (76%). Similarly, 77% of all respondents agreed to being comfortable speaking to their faith leader about their specific problems with no significant difference by gender (p=0.72) or region (p=0.09). With reference to respondents feeling a sense of connection to all life, slightly over a third (78%) of respondents agreed they felt connected to all life. There was no significant difference by gender (p=0.57) or region (p=0.10).

**Table 8:**
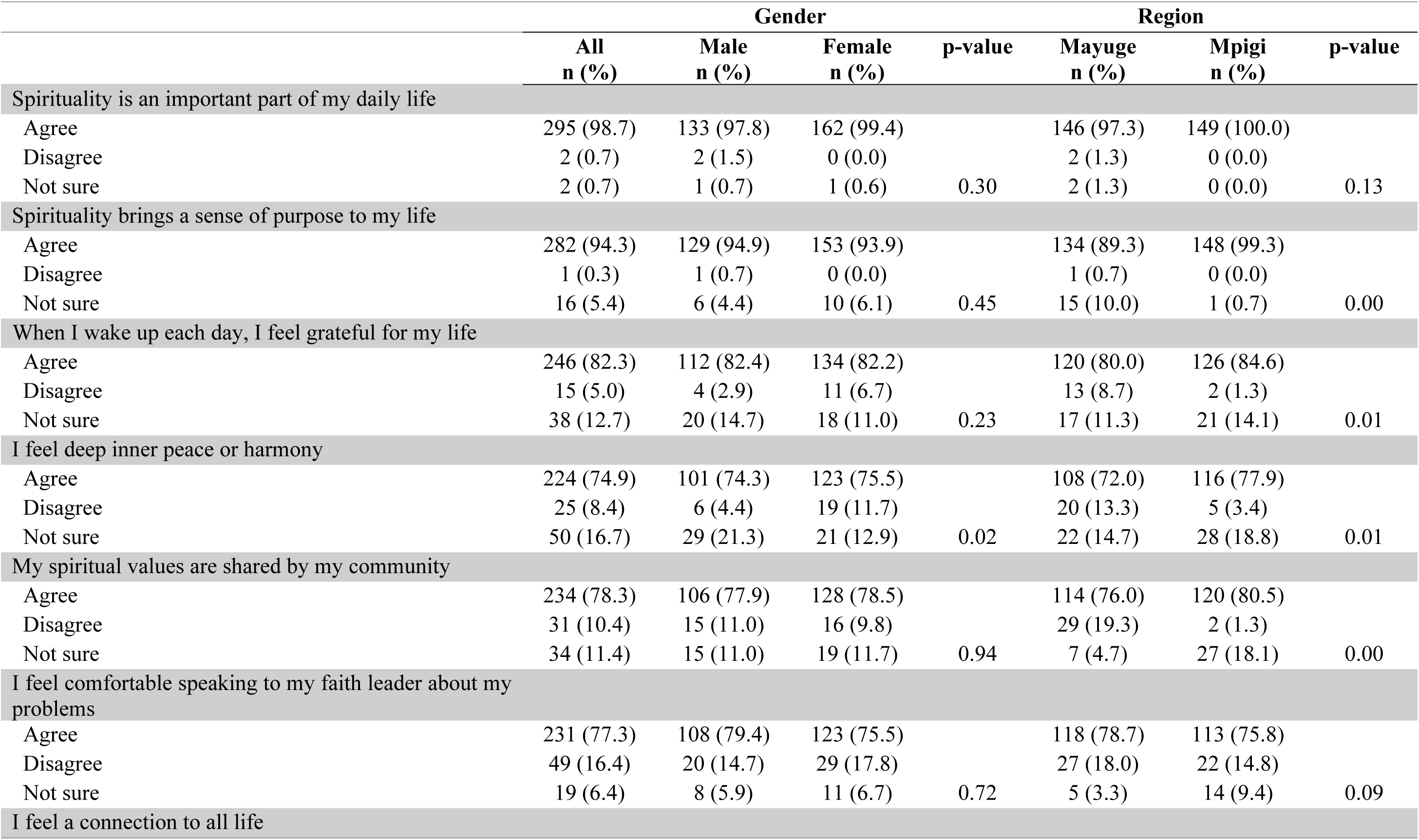
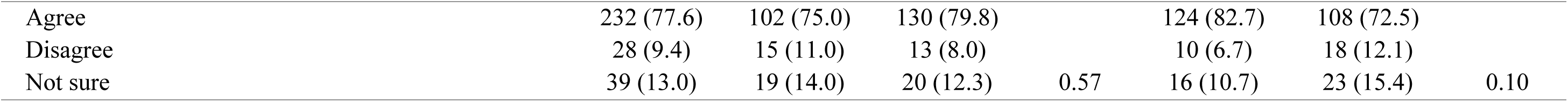
Theme Spiritual Purpose scores disaggregated by gender and region.

Almost all participants agreed that spirituality was an important part of their lives as it helps to bring a sense of purpose and inner peace as well as harmony to their life. Most of the respondents opined that their spiritual values are shared by the communities they dwell in. In addition, a majority of them reported being comfortable speaking to their faith leader about their specific problems. The findings therefore infer that within the community of Mayuge and Mpigi, spiritual purpose is acknowledged by the youth, and it plays a key role towards their flourishing in life. In other words, by using spiritual purpose, the youth can establish a set of principles, values and beliefs that give a sense of meaning to their life. In addition, this spiritual sense guides the decisions and actions they make.

### Equality Measure

Equality measures were assessed using a set of three indicators. They were inequalities in society, power to change inequalities and less inequalities in future. The results are presented in **Table 9**. Respondents were asked to score whether they saw a lot of inequalities in their respective communities and the majority (67%) scored on the affirmative with more being from Mayuge (70%) compared to Mpigi (64%). Respondents also scored their perceptions on whether they had the power to change inequalities in their communities in which only 40% reported they did, and this was higher in Mpigi (54%) compared to Mayuge (25%), p<0.001. Lastly, 80% of all respondents reported that they imagined a future life with less inequality. There was no significant difference in this by gender (p=0.45) or region (p=0.40).

**Table 9:** Theme; Equality Measures scores disaggregated by gender and region.

|  |  | Gender |  |  | Region |  |  |  |
| --- | --- | --- | --- | --- | --- | --- | --- | --- |
|  |  | All<br>n (%) | Male<br>n (%) | Female<br>n (%) | p-value | Mayuge<br>n (%) | Mpigi<br>n (%) | p-value |
| I see a lot of inequalities in my community |  |  |  |  |  |  |  |  |
| Agree |  | 200<br>(66.9) | 88 (64.7) | 112<br>(68.7) |  | 105<br>(70.0) | 95<br>(63.8) |  |
| Disagree |  | 54 (18.1) | 27 (19.9) | 27 (16.6) |  | 17 (11.3) | 37<br>(24.8) |  |
| Not sure |  | 45 (15.1) | 21 (15.4) | 24 (14.7) | 0.72 | 28 (18.7) | 17<br>(11.4) | 0.01 |
| I have the power to change inequalities in my community |  |  |  |  |  |  |  |  |
| Agree |  | 119<br>(39.8) | 56 (41.2) | 63 (38.7) |  | 38 (25.3) | 81<br>(54.4) |  |
| Disagree |  | 112<br>(37.5) | 50 (36.8) | 62 (38.0) |  | 70 (46.7) | 42<br>(28.2) |  |
| Not sure |  | 68 (22.7) | 30 (22.1) | 38 (23.3) | 0.90 | 42 (28.0) | 26<br>(17.4) | 0.00 |
| I imagine a future with less inequality |  |  |  |  |  |  |  |  |
| Agree |  | 240<br>(80.3) | 105<br>(77.2) | 135<br>(82.8) |  | 116<br>(77.3) | 124<br>(83.2) |  |
| Disagree |  | 31 (10.4) | 17 (12.5) | 14 (8.6) |  | 17 (11.3) | 14 (9.4) |  |
| Not sure |  | 28 (9.4) | 14 (10.3) | 14 (8.6) | 0.45 | 17 (11.3) | 11 (7.4) | 0.40 |

Evidently, the majority of the respondents reported that they saw a lot of inequalities in their respective communities. However, less than half of the respondents expressed confidence in changing inequalities in their communities. Finally, most of them imagined a future life with less inequality.

### Correlation Matrix

We examined relationships between different themes using correlation analysis by correlating composite scores generated from each theme. The results should however be interpreted with caution given the low internal consistency among the items used in each theme based on the Cronbach’s alpha coefficient (**Table 10**) for which it is recommended that Cronbach’s alpha coefficient of greater or equal to 7. Though the scores were <7, the findings can be used to show themes that potentially have high relationship. All themes at the individual theme level had alpha coefficient of <0.7, but the overall coefficient was 0.88.

**Table 10:** Reliability Statistics for each of the Themes Assessed (Cronbach Alpha)

|  | Number of items | Cronbach's Alpha | Remark |
| --- | --- | --- | --- |
| Knowledge/Education | 8 | 0.52 | Low |
| Emotional health | 14 | 0.65 | Low |
| Physical health | 8 | 0.52 | Low |
| Physical safety | 8 | 0.55 | Low |
| Financial security | 8 | 0.61 | Low |
| Social connectedness | 8 | 0.58 | Low |
| Spiritual purpose | 7 | 0.64 | Low |
| Equality measures | 3 | 0.36 | Low |
| Overall Reliability Test (All questions) | 64 | 0.88 | Adequate |

Figure 1 presents the findings. Deep blue cells represent correlation coefficients that were not statistically significant and red/pink cells represent cells that had positive significant correlation. The thicker the color (red/pink), the stronger the correlation. Interpretation of each individual correlation coefficient is based on the classification presented in **Table 1** under the methods section. While correlation analysis does not infer cause and effect relationship, it gives indication of associations among themes.

**Figure 1:**
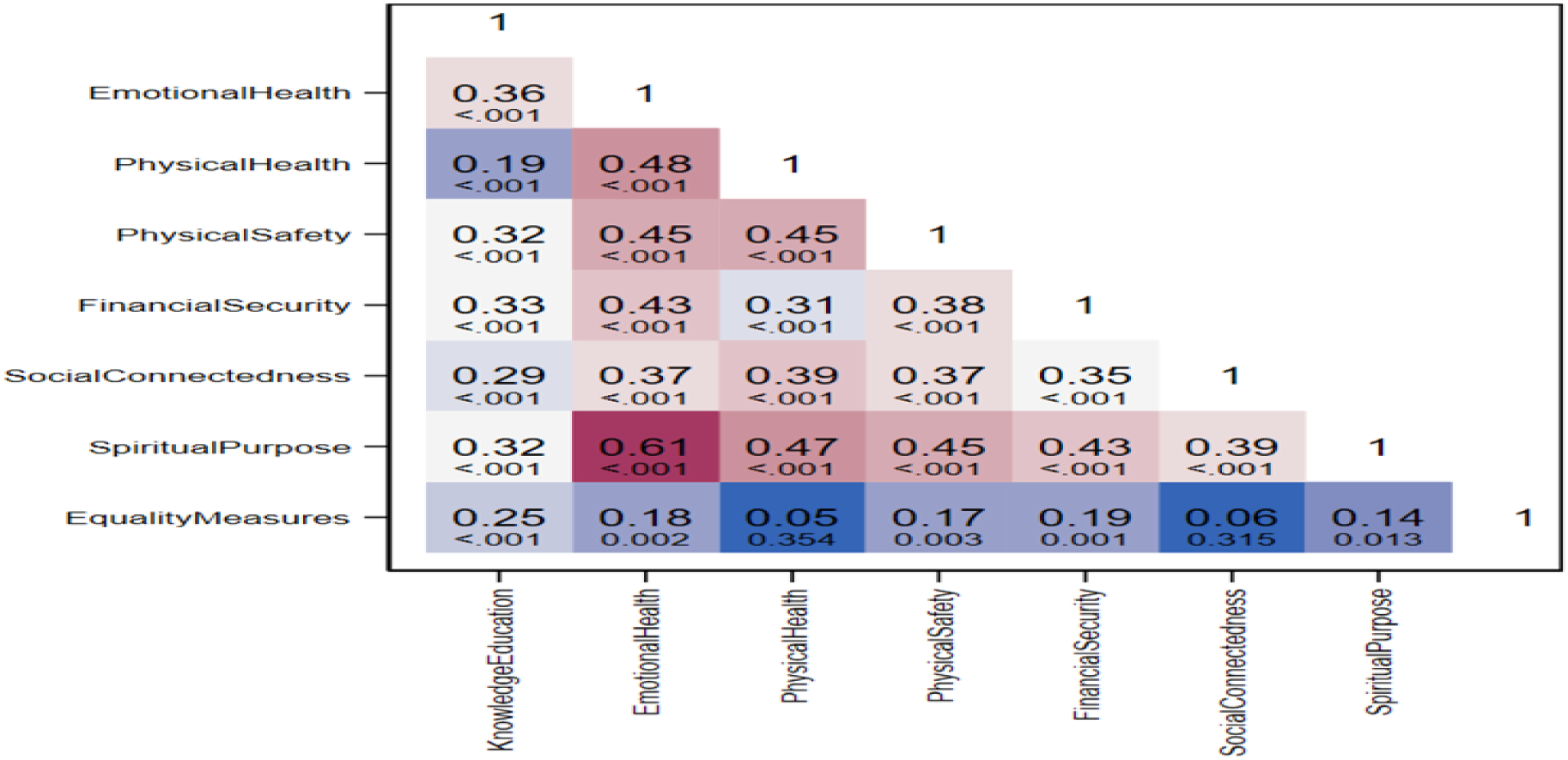
Correlation Matrix on the Relationship between the Thematic Areas Assessed.

The themes with the highest correlation included spiritual purpose and emotional health with a moderate positive correlation coefficient of 0.61, followed by emotional health and physical health with a moderate positive correlation coefficient of 0.48. Physical safety and emotional health had moderate positive correlation coefficient of 0.45, like physical health and physical safety. Spiritual purpose and physical health also had moderate positive correlation of 0.47 and 0.45 with physical safety. Correlation coefficients of less than 0.45 were classified as weak.

## 6. Discussions

In order to understand the determinants of flourishing among adolescents in Mayuge and Mpigi in Uganda, the study considered seven variables, namely, emotional health, financial security, physical safety, physical health, social connectedness, spiritual purpose and equality measure. Questions were formulated under each variable in consideration of some indicators, and they were subjected to the selected respondents. The respondents provided informed responses to each question. Furthermore, from a background perspective, the youth in Mayuge and Mpigi live in societies where they are faced with many inequalities; besides, very few can influence change. Nonetheless, they hope to live in a society with less inequalities in future.

As a result, the need for government intervention is apparent in areas like policy implementation that could reduce social inequalities in the country in general and within the two communities. Despite the enabling legal and policy framework in place, the Uganda Gender Policy (MGLSD, 2007a) acknowledges high incidences of early marriage, teenage pregnancy, and early sexual debut, all of which are violations of girls’ rights that have impacts on their health, the health of their children and future advancement.

In addition, the policy acknowledges persistent cultural norms and values that condone gender discrimination that have rendered the abuse of women’s rights socially acceptable in the Ugandan society. Under its ‘gender and rights’ priority area, the Gender Policy pledges to enact and reform laws to address gender-discriminatory practices, cultural norms and values; to develop and implement interventions to address GBV of all forms and at all levels; to promote sexual and reproductive health rights; and to sensitize communities about children’s rights; however, the strategies remain broad, with no specific explicit intervention to address early marriage (Bantebya, Muhanguzi, & Watson 2014). While acknowledging the importance of physical safety for their wellbeing and indicating that they felt secure both at home and at school, the key concern however is about the majority of young women respondents who felt unsafe due to their gender. Young women respondents therefore express their vulnerability because of their gender. This is a key aspect of psychological and emotional safety that needs to be considered for comprehensive physical safety of young people.

The government and the community should therefore ensure that policies and laws are put in place to address the concern of women feeling unsafe or vulnerable because of their gender. The government in general and the community members in particular need to put in place policies that guarantee women and girls their physical security. Therefore, policies should sustainably address issues pertaining to sexual and gender-based violence and violence against women within the communities.

Almost all respondents agreed that a balanced diet was important for their health and well-being. However, only about half of the respondents reported being able to decide on what they eat. Conversely, the analysis revealed that while the youth are aware of the importance of a balanced diet, they are not able to decide what to eat. On sanitation and hygiene, most of the respondents reported having access to clean water with most of them practicing basic hygiene. Some participants noted that they have health problems that prevented them from doing things that others of the same age could do; nevertheless, most of them were able to share their health concerns with their friends.

The respondents were also aware of safe sexual practices with a majority believing that safe sexual practices would allow them to have a brighter future. Moreover, while they are aware and believe in the importance of safe sex practices for brighter future, the study did not establish whether they did practice them. The study would conclude that it is difficult to establish whether the youth of Mayuge and Mpigi enjoy good physical health.

As far as financial security was concerned, the study revealed the inability of the respondents to meet their monthly living expenses, as a result, most of them reported feeling stressed. This means that even if they can secure a job that they like, the job is not sustainable to ensure financial security. This infers that financial insecurity is a big challenge facing the youth in Mayuge and Mpigi, a hindrance to their flourishing in life. This calls for alternative ways to ensure financial security within the society, including the use of alternative income generating activities.

In regard to social connectedness, the study findings revealed that most respondents have many close friends, this was most evident among male respondents with a majority saying that they are satisfied with their friendships and relationships and that they made other people happy. Still, a higher proportion of the youth interviewed pointed out that their friendships were important for their emotional well-being. In addition, most respondents mentioned that they felt connected to the world and that there was an adult in school whom they talked to about their problems. Lastly, most respondents indicated that they knew that their families loved them deeply. The analysis of these findings therefore confirms the element of social connectedness among the youth in Mayuge and Mpigi contributing to their flourishing. As a result of social connectedness, the youth can lower anxiety and depression, regulate their emotions, have higher self-esteem and empathy, and actually improve their immune systems.

Finally, spiritual purpose was acknowledged as an important aspect of youth lives as it helps to bring a sense of purpose, inner peace, and harmony. The spiritual values are shared among their communities further promoting harmonious living. Moreover, because of spiritual purpose, the youth can comfortably share their issues with their faith leaders. The findings therefore infer that within the community of Mayuge and Mpigi, spiritual purpose is acknowledged by the youth, and it plays a key role towards their flourishing in life. In other words, by using spiritual purpose, the youth can establish a set of principles, values and beliefs that give a sense of meaning to their life; therefore, guiding the decisions and actions that they make.

## 7. Conclusions and Recommendations

In conclusion, despite inequalities in their respective communities, the flourishing of youth in Mayuge and Mpigi is in good course as the majority confirmed that they are emotionally and physically healthy, physically safe, socially connected and have a spiritual purpose for life. However, the big challenge they are facing is related to their financial insecurity.

Little is known about the prevalence of mental disorders among Ugandan adolescents. This is a critical gap, as studies have found that mental disorders during adolescence, particularly those that remain untreated or under-treated, can have adverse outcomes throughout the life course. The literature that is available on adolescent mental disorders in Uganda is limited by small sample sizes, restricted locations or age ranges, a lack of diagnostic assessments (i.e., using symptom measures rather than trying to identify a disorder), and a focus on specific disorders. This means that the evidence to inform adolescent mental health policy in Uganda, and the subsequent ability to act positively for adolescent mental health, is extremely limited.

In consideration of the research findings, the following recommendations are made to the key stakeholders in general and to the youth of the two communities in particular:

● Adolescents who need sexual and reproductive health (SRH) services, should be supported by referring them to the Ministry of Health facilities that offer youth-friendly services. For the IS adolescents, the partners should work with parents/guardians to encourage them to hold sexuality talks with their children. Out-of-school activities (e.g., during holidays or weekends) that discuss sexuality topics should be recommended to the school going adolescents.
● A Holistic model should be developed to enhance the capacity of adolescents towards flourishing which will guide on the clear understanding of how physical, mental, spiritual, economic, cultural, and social dimensions of well-being intersect to express human flourishing. In addition, there is a need to use community engagement tools to address issues facing communities. For instance, the tools can be used to improve the perception of communities on gender norms, roles, stereotypes, and the effects of harmful traditional practices, as well as address the concerns that parents have regarding school adolescents engaging in income generating activities (IGA).
● To address mental health concerns highlighted in the communities, the partners should develop linkages with programs to be implemented on mental illness prevention, treatment, support, and care to the victims.
● Lack of school fees was cited as one of the reasons for poor school attendance and eventual dropout, showing that these communities are economically vulnerable; hence, leading to mental issues of the adolescents. A follow-up intervention should also include communities as beneficiaries of the economic empowerment package.
● The government of Uganda and the communities should consider developing policies and measures to foster equality measures towards reducing inequalities within the communities.
● There is a need for the youth to seek alternative sources of income to assure financial security and sustainability. The communities, teachers, mentors, and parents of the youth should encourage the youth to take up income generating activities. They should also be supported financially to build the capital they need to flourish in business and entrepreneurship.
● The government and the communities should put in place policies and measures that will sustainably address GBV, SGBV and any other concern to ensure the physical security of women and girls who feel unsafe because of their gender.
● Enforce safety measures in the community: there is a need to carry out sensitization programs through holistic models and empowerment for adolescents and parents to curb teen pregnancies and child marriages.

## Data Availability

We have data protected and in store that can be available when requested

## Acknowledgements

Dr. Benson Mutuku, Dr. Carol Henry and Dr. Judy White wrote this report. Several individuals from different institutions contributed to the study and review of the report. Dr. Benson Mutuku in particular, a gender expert, steered the work beginning with a comprehensive process of data cleaning and analysis. Together with the other team members, we held consultation with diverse stakeholders, development of the study protocol, review of the study tools, and the review process for all reports. The other team members from partner institutions who supported throughout the process are Dr, Benjamin Lough, Dr. Rebecca Tiessen, Dr. Daniel Kikulwe, Dr. Asrat Tolossa, Melani O’Leary (formerly World Vision, Canada), Sarah Crawford, Dominic Schofield, James Kimmula, Victor O Hardy, Lydia Kabiri and Dr. David Musoke.

We are indebted to teams from different partner institutions (University of Saskatchewan, World Vision Canada and Uganda, Universities of Regina, Illinois, and Ottawa, York University, and Makerere University) who provided robust leadership and guidance throughout the whole process. The Ugandan team facilitated the identification and engagement with the focal point persons who collected the primary data and reviewed the reports with a contextual lens. The team from Canada reviewed the report from a technical perspective. A final and special acknowledgement goes to the participants who volunteered to participate in the study, and the enumerators who offered valuable support in the interview process. The in-country mobilizers were instrumental in coordinating this process. The team wishes to thank the Makerere University School of Health Sciences Institutional Review Board for granting clearance for the research.

This research was supported by the World Charity Foundation, USA, and the Global Institute for Food Security, University of Saskatchewan, Canada.

## Disclaimer

*The views expressed in this document are those of the authors and do not necessarily reflect the views of Templeton Foundation, or other institutions involved*.

## Notes

### Competing Interest Statement

The authors have declared no competing interest.

### Funding Statement

Funding for this research was provided by Templeton foundation Canada and University of Saskatchewan

### Author Declarations

The University of Saskatchewan Behavioral Research Ethics Board (Beh-REB)

